# Legal residency status and its relationship with health indicators among Syrian refugees in Lebanon: a nested cross-sectional study

**DOI:** 10.1101/2024.09.27.24314488

**Authors:** Marie-Elizabeth Ragi, Hala Ghattas, Berthe Abi Zeid, Hazar Shamas, Noura Salibi, Sawsan Abdulrahim, Jocelyn DeJong, Stephen J. McCall, CAEP study group

**Affiliations:** Center for Research on Population and Health, Faculty of Health Sciences, American University of Beirut, Beirut, Lebanon; Department of Health Promotion, Education, and Behavior, University of South Carolina, Columbia, South Carolina, USA; Department of Health Promotion and Community Health, Faculty of Health Sciences, American University of Beirut, Beirut, Lebanon; Department of Epidemiology and Population Health, Faculty of Health Sciences, American University of Beirut, Beirut, Lebanon

**Author notes:** Corresponding author: Stephen J McCall, Center for Research on Population and Health, Faculty of Health Sciences, American University of Beirut, Beirut 1107 2020, Lebanon.

**Keywords:** Syrian refugees, Legal residency, Health outcomes

## Abstract

**Background:** Failure to possess or renew legal residency permits increases the burden on a vulnerable refugee population. It risks detention or deportation, and hinders access to basic services including healthcare. This study aimed to examined the association between legal residency status and health of Syrian refugees living in Lebanon.

**Methods:** Data were from two independent nested cross-sectional studies collected in 2022 through telephone surveys. In the first study, all Syrian refugees aged 50 years or older from households that received humanitarian assistance were invited to participate. The second included all adult Syrian refugees residing in a suburb of Beirut. The exposure was self-reported possession of a legal residency permit in Lebanon. The self-reported health outcomes were mental health status, COVID-19 vaccine uptake, and access to the needed healthcare services. Separate adjusted logistic regression models examined the association between lacking a legal residency permit and each health outcome.

**Results:** The first sample included 3357 participants (median age 58 years (IQR:54-64), 47% female), of whom 85% reported lacking a legal residency permit. The second sample included 730 participants (median age 34 years (IQR:26-42), 49% female), of whom 79% lacked a legal residency permit. In both studies, lacking a legal residency permit increased the odds of having poor mental health [adjusted odds ratio (aOR):1.62 (95%CI:1.2-2.2); aOR:1.62 (95%CI:1.01-2.60)], and decreased the odds of COVID-19 vaccine uptake [aOR:0.64 (95%CI:0.53-0.78); aOR:0.51 (95%CI:0.32-0.81)]. In the sub-sample who needed primary healthcare, lacking a legal residency permit decreased the odds of access to primary healthcare in the second study only (aOR:0.37 (95%CI:0.17-0.84)).

**Conclusions:** The majority of Syrian refugees from these two samples reported lacking a legal residency permit in Lebanon. This was associated with poor mental health and lower uptake of COVID-19 vaccination, potentially originating from fear of detention or deportation. These findings call for urgent action to support access to legal documentation for refugees in Lebanon.

**Key Messages:** *What is already known on this topic:* Refugees are a vulnerable population and face varied challenges, such as marginalization and high levels of poverty. The lack of legal residency increases the risk of detention or deportation and may impact access to essential public services and healthcare.

*What this study adds:* This study showed that the majority of Syrian refugees in Lebanon lacked legal residency permits and this impacted receipt of the COVID-19 vaccination during the pandemic and their mental health.

*How this study might affect research, practice or policy:* These findings highlight the need for actions to support access for legal documentation for refugees and enable equitable access to vaccination campaigns and health and mental health services for this vulnerable population.

## Introduction

Following the onset of the Syrian civil war in 2011, large numbers of Syrians were displaced and sought refuge in neighbouring countries, with Lebanon, Jordan and Turkey being the primary host countries for Syrian refugees [1]. Currently, Lebanon hosts the world’s highest per capita population of refugees, with over 1.5 million Syrian refugees estimated by the Lebanese government [2], including about 815,000 registered with UNHCR [3]. Many live in urban residential areas among Lebanese communities, while about 20% live in informal tented settlements [2, 4].

Refugee populations are particularly vulnerable to exclusion, stigma and discrimination driven by social attitudes and official legislation [5–7]. They face multiple challenges such as restricted mobility [2, 8, 9], high levels of poverty and crowding [2, 7], and inadequate access to education, healthcare, and other services [2, 10–12]. Barriers to adequate healthcare access for Syrian refugees in Lebanon have been reported to include access to transportation and economic resources, as well as discrimination [13]. Discrimination itself is a determinant of poor mental and physical health among Syrian refugees in Turkey [14].

In addition, the Lebanese government requires Syrian nationals to enter Lebanon legally and renew their legal residency permits annually. This significantly burdens Syrian refugee households due to high cost of renewal and paperwork requirements [4, 15, 16]. Recent data indicate that ~80% of Syrians residing in Lebanon do not hold legal residency permits [2, 4, 17]. Failure to possess a valid residency permit in the country increases the risk of detention, deportation, or exposure to abuse or exploitation, thus adding to the multifaceted strains on Syrian refugees [4, 15, 16, 18]. This limits freedom of movement, generates barriers to access work permits and public services, including healthcare, and exacerbates disparities of protracted displacement [16–20]. Studies globally highlighted the critical impact of refugees’ legal status on healthcare access, reporting that refugees lacking legal documentation are excluded from the national healthcare systems and restricted to emergency care thus leading to health disparities, and detrimental physical and mental health outcomes [21–23]. In Lebanon, for example, most healthcare facilities require refugees to present identification, which poses challenges to those without documentation [24] and potentially hinders healthcare access due to fear of deportation [25].

Yet, the literature remains limited in comparing health outcomes between those with and without a legal residency permit. Consequently, the impact of legal residency status on the multiple health challenges faced by Syrian refugees remains poorly understood. Thus, we examine the association between the lack of a legal residency permit and several health indicators among two sub-populations of Syrian refugees living in Lebanon.

## Methods

### Study design and setting

We conducted a nested cross-sectional analysis from two multi-wave longitudinal surveys, both of which aimed to track the vulnerabilities of Syrian refugees in the context of the COVID-19 pandemic in Lebanon. The first (Changing Vulnerabilities and COVID adherence – CVC study) included Syrian refugees aged 50 years or older from different areas of Lebanon. The second (Community Action for Equity in Pandemic preparedness and control – CAEP study) included all adult Syrian refugees aged 18 years and older residing in Sin-El-Fil, a suburb of Beirut, Lebanon. Sin El Fil hosts a large number of migrants and refugees who reside closely alongside the Lebanese host community, while it also has a diverse range of socio-economic levels.

### Sampling and study population

The CVC Study sampling frame included all households in Lebanon that received assistance between 2017 and 2020 from the Norwegian Refugee Council, a humanitarian non-governmental organization (NGO), and which comprised Syrian adults aged 50 years or older (n=17,384). All households in the sample listing were contacted, and Syrian refugees aged 50 years or older were invited to participate in the study. If households had more than one older adult, one older adult was randomly selected by a computer algorithm when a household was included to participate. This analysis used data from participants who completed the fifth wave of data collection from January to March 2022 (n=3,370) (Supplementary Figure 1).

The CAEP study followed a multi-stage stratified sampling design using an area-based sampling approach [26]. A household listing exercise was carried out in April 2022, whereby all households in Sin-El-Fil were enumerated face-to-face to complete an eligibility screening survey, which identified all households that included adults with Syrian nationality [27]. Pre-consent was obtained from all households with eligible individuals, and phone numbers were collected from consenting households. For the study sample selection, a listing of all eligible household members was generated. All Syrian adults were selected and included in the study. For the present analysis, the study population included all Syrian participants who completed the first wave of data collection conducted between June and October 2022 (n=739) (Supplementary Figure 2).

For both studies, respondents were contacted to complete a computer-assisted telephone survey conducted by a trained data collector using KoboToolbox (CVC study) and SurveyCTO (CAEP study) software (Cambridge, MA, USA). Verbal informed consent to participate in a telephone interview was obtained from all participants, and those aged 65 (CVC study) or 60 (CAEP study) years or older were further assessed for capacity to consent using 5 modified items from the University of California, San Diego, Brief Assessment of Capacity to Consent [28].

### Data sources

For the CVC study, the questionnaire included existing validated scales and contextualised questions. It was co-created by academics, humanitarian workers, government representatives, and focal points from the refugee community. The survey tool was drafted in English and then translated into Arabic. Before data collection, the Arabic version of the questionnaire was piloted internally with data collectors and local community focal points for face validity. Several adjustments to the survey instrument were conducted on the basis of the pilot test, community consultations, and data collectors’ training accordingly. Data were monitored daily in parallel with data collection for quality assurance. Details on the development of the survey tool have been described elsewhere [29, 30].

The questionnaire for the CAEP study was developed using existing questionnaire modules, validated scales, contextualized questions, and community-identified priorities. The survey was created in collaboration with representatives from the municipality of Sin-El-Fil, the Ministry of Public Health (MOPH), and NGOs operating in Sin-El-Fil. The survey tool was drafted in English, translated to Arabic and tested for comprehension prior to deployment. Data were frequently monitored in parallel with data collection for quality assurance, with call back checks conducted for verification purposes on 5% of the sample.

Both studies primarily used similar questions and comparable variables for data harmonization purposes. A mapping exercise was carried out to ensure matched coding, and in case of divergences, harmonised coding and categorization were generated.

### Exposure and Outcomes measures

The exposure was self-reported possession of a legal residency permit in Lebanon (yes/no). In both studies, each participant was asked the following question: “Do you have regularized residency in Lebanon?”. Possession of a valid residency permit, obtained by a yearly renewal of residency permits, is indicative of legal residency or regularized residency in Lebanon.

The health outcomes were self-reported. They included for the CVC study mental health status defined as poor with a Mental Health Inventory-5 score of less or equal to 60, uptake of at least one dose of the COVID-19 vaccine (yes/no), access to primary health care services for those who needed primary healthcare in the previous month (yes/no), and chronic disease management in those who reported being diagnosed with a chronic disease (yes/no). For the CAEP study, the health outcomes were mental health status defined using Patient Health Questionnaire-9 (PHQ-9) and General Anxiety Disorder-7 (GAD-7) scores of less than 10 indicating moderate to severe depression or anxiety respectively, uptake of at least of dose of the COVID-19 vaccine (yes/no), access to primary health care services for those who needed primary healthcare in the previous month (yes/no), and medication access for those who were prescribed medication for the management of diagnosed chronic diseases (yes/no). In both studies, socio-demographic characteristics were also collected and included age (continuous), sex (male/female), marital status (single or engaged/ married/ separated or widowed), length of stay in Lebanon (continuous), educational level (never attended school/ attended school), current employment status (yes/no), wealth index (lowest/ middle/ highest tertile), and receipt of assistance (yes/no). Residence inside or outside Informal Tented Settlements (ITS) was reported for the CVC study sample. The household assets-based wealth index was generated through Principal Component Analysis (PCA) using household assets that includes transportation, communication, home technology and cooking assets. Missing values were missing at random [31], all variables had missing values of less than 7.5% and complete case analysis was performed [32].

### Statistical analysis

Frequencies and percentages were calculated for categorical variables, and medians and Interquartile Ranges (IQRs) for continuous variables. Pearson’s chi-squared test and logistic regression were used to assess differences between those with and without a legal residency permit for categorial and continuous variables respectively. For the CAEP study, the analysis was weighted according to the sampling selection probabilities and non-response; the study weight was generated from the product of the probability of selection adjustment in the study sample and the non-response adjustment computed by using a response propensity logistic regression model to account for attrition in wave 1 of data collection [33]. The exposure was a categorical variable on the possession of a legal residency permit and all health outcome variables were binary. The association between legal residency and the different outcomes for each study samples was examined in separate analyses using bivariate logistic regression analyses, and unadjusted odds ratios (ORs) along with their 95% confidence intervals (CIs) are reported. Within each study sample, logistic regression models were used to estimate the association between the lack of a legal residency permit and each outcome, adjusted for age, length of stay in Lebanon, education, employment, wealth index, and receipt of assistance which were identified using Directed Acyclic Graphs (Supplementary Figure 3). A p-value less than 0.05 was considered statistically significant. All analyses were conducted using Stata/SE statistical software version 18 (STATACorp).

### Ethical approval and reporting

This study was reported according to the STROBE reporting guidelines [34]. Both study protocols were reviewed and approved by the American University of Beirut Social and Behavioral Sciences Institutional Review Board (reference: SBS-2020-0329 for the CVC study & SBS-2021-0268 for the CAEP study).

## Results

### CVC study descriptive

A total of 3,357 participants who completed the fifth wave of data collection and reported their legal residency status were included in the analysis. Median age was 58 years (IQR 54-64) and 47.3% were female (Table 1). From the CVC study sample, 85.3% (n=2,864) reported lacking a legal residency permit in Lebanon. Differences were observed in socio-demographic indicators between those without and with a legal residency permit (Table 1), where participants without a legal residency permit had higher median age (59- vs. 58-years old, p=0.005), lower median length of stay in Lebanon (7- vs. 8-years, p=0.008), a higher proportion of females (48.5% vs. 40.4%, p=0.001), lower educational level (49.5% vs. 40.6% having never attended school, p<0.001), poorer wealth (40.0% vs. 28.2% in the lowest tertile of wealth index, p<0.001), and lower employment (4.7% vs. 7.1%, p=0.023). The majority of the sample, with more than 90% in both those without and with a legal residency permit, reported receiving cash assistance; and no significant difference in legal residency status was observed between those living inside vs outside ITS.

**Table 1.** CVC study – Characteristics of the older adult Syrian refugee population from all areas of Lebanon by possession of a legal residency permit.

|  | **Total**<br>**n=3,357** |  | **No legal residency**<br>**permit**<br>**n=2,864 (85.3%)** |  | **Possession of a legal**<br>**residency permit**<br>**n=493 (14.7%)** |  | **P-value** |
| --- | --- | --- | --- | --- | --- | --- | --- |
|  | **n** | **%** | **n** | **%** | **n** | **%** |  |
| **Age [years] median (IQR)** | 3,357 | 58 (54-64) | 2,864 | 59 (55-65) | 493 | 58 (54-63) | 0.005 |
| **Length of stay in Lebanon median (IQR)** | 3,328 | 7 (7-8) | 2,840 | 7 (6-8) | 488 | 8 (7-8) | 0.008 |
| Missing | 29 |  | 24 |  | 5 |  |  |
| **Sex** |  |  |  |  |  |  |  |
| Male | 1,768 | (52.7) | 1,474 | (51.5) | 294 | (59.6) | 0.001 |
| Female | 1,589 | (47.3) | 1,390 | (48.5) | 199 | (40.4) |  |
| **Marital status** |  |  |  |  |  |  |  |
| Single/Engaged | 29 | (0.9) | 27 | (1.0) | 2 | (0.4) | 0.005 |
| Married | 2,382 | (71) | 1,997 | (69.7) | 385 | (78.1) |  |
| Separated/Widowed | 946 | (28.2) | 840 | (29.3) | 106 | (21.5) |  |
| **Education** |  |  |  |  |  |  |  |
| Never attended school | 1,613 | (48.2) | 1,414 | (49.5) | 199 | (40.6) | <0.001 |
| Attended school | 1,735 | (51.8) | 1,444 | (50.5) | 291 | (59.4) |  |
| Missing | 9 |  | 6 |  | 3 |  |  |
| **Employment** |  |  |  |  |  |  |  |
| Currently not employed | 3,188 | (95.0) | 2,730 | (95.3) | 458 | (92.9) | 0.023 |
| Currently employed | 169 | (5.0) | 134 | (4.7) | 35 | (7.1) |  |
| **Residence inside/outside ITS** |  |  |  |  |  |  |  |
| Inside ITS | 1,262 | (37.6) | 1,087 | (37.9) | 175 | (35.5) | 0.298 |
| Outside ITS | 2,095 | (62.4) | 1,777 | (62.1) | 318 | (64.5) |  |
| **Wealth index** |  |  |  |  |  |  |  |
| Lowest | 1,286 | (38.3) | 1,147 | (40.0) | 139 | (28.2) | <0.001 |
| Middle | 1,014 | (30.2) | 872 | (30.5) | 142 | (28.8) |  |
| Highest | 1,057 | (31.5) | 845 | (29.5) | 212 | (43.0) |  |
| **Receipt of cash assistance** |  |  |  |  |  |  |  |
| No | 247 | (7.4) | 204 | (7.1) | 43 | (8.8) | 0.126 |
| Yes | 3,101 | (92.6) | 2,654 | (92.9) | 447 | (91.2) |  |
| Missing | 9 |  | 6 |  | 3 |  |  |
| **Mental Health Score** |  |  |  |  |  |  |  |
| Good | 306 | (9.1) | 243 | (8.5) | 63 | (12.8) | 0.002 |
| Poor | 3,051 | (90.9) | 2,621 | (91.5) | 430 | (87.2) |  |
| **COVID-19 vaccine uptake** |  |  |  |  |  |  |  |
| Not vaccinated | 1,927 | (57.5) | 1,692 | (59.2) | 235 | (47.9) | <0.001 |
| Vaccinated with at least one dose | 1,422 | (42.5) | 1,166 | (40.8) | 256 | (52.1) |  |
| Missing | 8 |  | 6 |  | 2 |  |  |
| **Primary healthcare access among those who needed healthcare** |  |  |  |  |  |  |  |
| Not able to access | 647 | (41.8) | 558 | (42.6) | 89 | (37.4) | 0.135 |
| Able to access | 901 | (58.2) | 752 | (57.4) | 149 | (62.6) |  |
| Missing | 1 |  | 0 |  | 1 |  |  |
| **Management of chronic diseases among those with chronic diseases** |  |  |  |  |  |  |  |
| Not able to manage | 374 | (16.9) | 325 | (17.3) | 49 | (14.6) | 0.230 |
| Able to manage | 1,840 | (83.1) | 1,554 | (82.7) | 286 | (85.4) |  |

*Data are n and % or median (IQR), IQR= Interquartile range.*
*Age and Length of stay Lebanon are continuous variables.*

### CAEP study descriptive

A total of 730 participants who reported their legal residency status were included in the analysis. The median age was 34 years old (IQR 26-42) and 49.4% were female (Table 2). In this sample, 79.4% (n=584) reported lacking a legal residency permit in Lebanon. No age and gender differences were observed between those without and with legal residency permit (p=0.777 and p=0.241, respectively). Those without a legal residency permit (Table 2) had lower median years of living in Lebanon (9- vs. 10-years, p<0.001), poorer wealth (81.8% vs. 56.8% in the lowest tertile of wealth, p<0.001), higher receipt of assistance (51.9% vs. 31.1%, p<0.001), and lower employment (37.1% vs. 49.2%, p=0.013).

**Table 2.** CAEP study - Characteristics of adult Syrian population living in Sin El Fil – Lebanon by possession of a legal residency permit.

|  | Total<br>n=730 |  | No legal residency<br>permit<br>n=584 (79.4%) |  | Possession of a legal<br>residency permit<br>n=146 (20.6%) |  | P-value |
| --- | --- | --- | --- | --- | --- | --- | --- |
|  | n | Weighted<br>% | n | Weighted<br>% | n | Weighted<br>% |  |
| **Age [years] median (IQR)** | 728 | 34 (26-42) | 582 | 35 (25-41) | 146 | 34 (26-42) | 0.777 |
| Missing | 2 |  | 2 |  | 0 |  |  |
| **Length of stay in Lebanon**<br>**median (IQR)** | 707 | 9 (7-11) | 566 | 9 (7-10) | 141 | 10 (8-17) | <0.001 |
| Missing | 23 |  | 18 |  | 5 |  |  |
| **Sex** |  |  |  |  |  |  |  |
| Male | 373 | (50.6) | 287 | (49.4) | 86 | (55.3) | 0.241 |
| Female | 357 | (49.4) | 297 | (50.6) | 60 | (44.7) |  |
| **Marital status** |  |  |  |  |  |  |  |
| Single/Engaged | 127 | (17.5) | 100 | (17.1) | 27 | (19.0) | 0.325 |
| Married | 570 | (77.7) | 455 | (77.5) | 115 | (78.5) |  |
| Separated/Widowed | 33 | (4.8) | 29 | (5.4) | 4 | (2.4) |  |
| **Education** |  |  |  |  |  |  |  |
| Never attended school | 108 | (15.2) | 90 | (15.5) | 18 | (14.3) | 0.757 |
| Attended school | 603 | (84.8) | 488 | (84.5) | 115 | (85.7) |  |
| Missing | 19 |  | 6 |  | 13 |  |  |
| **Employment** |  |  |  |  |  |  |  |
| Currently not employed | 432 | (60.4) | 363 | (62.9) | 69 | (50.8) | 0.013 |
| Currently employed | 298 | (39.6) | 221 | (37.1) | 77 | (49.2) |  |
| **Wealth index** |  |  |  |  |  |  |  |
| Lowest | 563 | (76.7) | 479 | (81.8) | 84 | (56.8) | <0.001 |
| Middle | 141 | (19.2) | 93 | (15.9) | 48 | (31.9) |  |
| Highest | 25 | (4.2) | 12 | (2.3) | 13 | (11.4) |  |
| Missing | 1 |  | 0 |  | 1 |  |  |
| **Receipt of assistance** |  |  |  |  |  |  |  |
| No | 366 | (52.3) | 274 | (48.1) | 92 | (68.9) | <0.001 |
| Yes | 344 | (47.7) | 295 | (51.9) | 49 | (31.1) |  |
| Missing | 20 |  | 15 |  | 5 |  |  |
| **Depression score PHQ-9** |  |  |  |  |  |  |  |
| None-Minimal/Mild | 410 | (57.0) | 311 | (53.6) | 99 | (70.3) | 0.001 |
| Moderate-Severe depression | 313 | (43.0) | 268 | (46.4) | 45 | (29.7) |  |
| Missing | 7 |  | 5 |  | 2 |  |  |
| **Anxiety score GAD-7** |  |  |  |  |  |  |  |
| Minimal/Mild | 373 | (52.8) | 290 | (51.2) | 83 | (58.9) | 0.118 |
| Moderate/Severe anxiety | 354 | (47.2) | 292 | (48.8) | 62 | (41.1) |  |
| Missing | 3 |  | 2 |  | 1 |  |  |
| **COVID-19 vaccine uptake** |  |  |  |  |  |  |  |
| Not vaccinated | 396 | (53.4) | 339 | (57.2) | 57 | (39.0) | <0.001 |
| Vaccinated with at least one dose | 329 | (46.6) | 240 | (42.8) | 89 | (61.0) |  |
| Missing | 5 |  | 5 |  | 0 |  |  |
| **Primary healthcare access among those who needed healthcare** |  |  |  |  |  |  |  |
| Not able to access | 95 | (46.5) | 82 | (51.0) | 13 | (31.3) | 0.037 |
| Able to access | 105 | (53.5) | 75 | (49.0) | 30 | (68.7) |  |
| **Medication access among those with chronic diseases** |  |  |  |  |  |  |  |
| Not taking at least one prescribed med | 35 | (29.3) | 31 | (29.8) | 4 | (26.6) | 0.816 |
| Taking all prescribed meds | 84 | (70.7) | 71 | (70.2) | 13 | (73.4) |  |

*PHQ-9 = Patient Health Questionnaire-9, GAD-7 = General Anxiety Disorder-7.*
*Data are n and weighted % or median (IQR), IQR= Interquartile range.*
*Age and Length of stay Lebanon are continuous variables.*

In both study populations, as compared to those with a legal residency permit, those lacking a legal residency permit had poorer mental health (91.5% vs. 87.2% had poor mental health (p=0.002) in the CVC study (Table 1); 46.4% vs. 29.7% had higher moderate to severe depression (p=0.001) in the CAEP study (Table 2)), and lower COVID-19 vaccination uptake (40.8% vs. 52.1% (p<0.001) in the CVC study; 42.8% vs. 61.0% (p<0.001) in the CAEP study). In the adjusted logistic regression, the lack of a legal residency permit in Lebanon increased the odds of having poor mental health (adjusted odds ratio (aOR):1.62 (95%CI:1.2-2.2)) in the CVC study population (Table 3) and moderate to severe depression (aOR:1.62 (95%CI:1.01-2.60)) in the CAEP study sample (Table 4). In addition, lacking a legal residency permit decreased the odds of having received a COVID-19 vaccine in both populations (CVC study: aOR:0.64 (95%CI:0.53-0.78) and CAEP study: aOR:0.51 (95%CI:0.32-0.81)).

**Table 3.** CVC study - Association between legal residency status and health outcomes in older adult Syrian refugee population from all areas of Lebanon.

|  | Unadjusted<br>logistic<br>regression | P-value | Adjusted<br>logistic<br>regression | P-value |
| --- | --- | --- | --- | --- |
|  | Weighted<br>OR [95% CI] |  | Weighted<br>OR [95% CI] |  |
| **Poor Mental Health Inventory-5 score** |  |  |  |  |
| Possession of a legal residency permit | 1.00 |  | 1.00 |  |
| No legal residency permit | 1.58 [1.18-2.12] | 0.002 | 1.46 [1.07 - 1.99] | 0.018 |
| **COVID-19 vaccine uptake** |  |  |  |  |
| Possession of a legal residency permit | 1.00 |  | 1.00 |  |
| No legal residency permit | 0.63 [0.52-0.77] | <0.001 | 0.66 [0.54 -0.80] | <0.001 |
| **Ability to access primary healthcare among those who needed healthcare** |  |  |  |  |
| Possession of a legal residency permit | 1.00 |  | 1.00 |  |
| No legal residency permit | 0.80 [0.61-1.07] | 0.135 | 0.83 [0.62 - 1.12] | 0.226 |
| **Ability to manage chronic diseases among those with chronic diseases** |  |  |  |  |
| Possession of a legal residency permit | 1.00 |  | 1.00 |  |
| No legal residency permit | 0.82 [0.59-1.13] | 0.230 | 0.83 [0.59 - 1.17] | 0.287 |

*OR=odds ratio. CI=confidence interval.*
*All outcomes adjusted for age, length of stay in Lebanon, education, employment, receipt of assistance, and wealth index according to the DAGs.*
*Employment (0=Currently not employed, 1=Currently employed); Mental Health Inventory-5 score (0=Poor, 1=Good); COVID-19 vaccine uptake (0=Not vaccinated, 1=Vaccinated with at least one dose); Primary healthcare access for those who required healthcare (0=Did not received the required care, 1=Received the required care); Chronic disease management in those who reporting being diagnosed with a chronic disease (0=Not able to manage, 1=Able to manage).*

**Table 4.** CAEP study - Association between legal residency status and health outcomes in adult Syrian population living in Sin El Fil, Lebanon.

|  | Unadjusted<br>logistic<br>regression | P-value | Adjusted<br>logistic<br>regression | P-value |
| --- | --- | --- | --- | --- |
|  | Weighted<br>OR [95% CI] |  | Weighted<br>OR [95% CI] |  |
| **Moderate to Severe Depression PHQ-9 score** |  |  |  |  |
| Possession of a legal residency permit | 1.00 |  | 1.00 |  |
| No legal residency permit | 2.05 [1.35-3.10] | 0.001 | 1.62 [1.01-2.60] | 0.045 |
| **Moderate to Severe Anxiety GAD-7 score** |  |  |  |  |
| Possession of a legal residency permit | 1.00 |  | 1.00 |  |
| No legal residency permit | 1.37 [0.92-2.02] | 0.119 | 1.04 [0.66-1.62] | 0.876 |
| **COVID-19 vaccine uptake** |  |  |  |  |
| Possession of a legal residency permit | 1.00 |  | 1.00 |  |
| No legal residency permit | 0.48 [0.32-0.71] | <0.001 | 0.51 [0.32-0.81] | 0.005 |
| **Ability to access primary healthcare among those who needed healthcare** |  |  |  |  |
| Possession of a legal residency permit | 1.00 |  | 1.00 |  |
| No legal residency permit | 0.44 [0.20-0.96] | 0.039 | 0.37 [0.17-0.84] | 0.017 |
| **Ability to access prescribed medication among those with chronic diseases** |  |  |  |  |
| Possession of a legal residency permit | 1.00 |  | 1.00 |  |
| No legal residency permit | 0.85 [0.22-3.24] | 0.816 | 1.01 [0.26-3.97] | 0.984 |

OR=odds ratio. CI=confidence interval.
PHQ-9 = Patient Health Questionnaire-9, GAD-7 = General Anxiety Disorder-7.
All outcomes adjusted for age, length of stay in Lebanon, education, employment, receipt of assistance, and wealth index according to the DAGs.
Employment (0=Currently not employed, 1=Currently employed); PHQ-9 (0=None-Minimal/Mild Depression, 1=Moderate to Severe Depression); GAD-7 (0=Minimal/Mild Anxiety, 1=Moderate/Severe Anxiety); COVID-19 vaccine uptake (0=Not vaccinated, 1=Vaccinated with at least one dose); Primary healthcare access for those who required healthcare (0=Did not received the required care, 1=Received the required care); Medication access in those who are prescribed medication for the management of diagnosed chronic disease (0=Not taking at least one prescribed medication, 1=Taking all prescribed medication).

In the sub-sample of participants who needed primary healthcare in the previous month (46% (n=1,549) in the CVC study and 27% (n=200) in the CAEP study), 42% (n=647) in the CVC study and 46% (n=95) in the CAEP study could not receive the needed healthcare. Differences by legal residency status were observed where a greater proportion of those without a legal residency permit were unable to access the needed healthcare services as opposed to those with legal residency for the CAEP study (51.0% vs. 31.3%, p=0.037) (Table 2), as well as the CVC study although it did not reach statistical significance (42.6% vs. 37.4%, p=0.135) (Table 1).Accordingly, those lacking a legal residency permit were found to have lower odds of access to primary healthcare (aOR:0.80 (95%CI:0.60-1.07) for the CVC study and aOR:0.37 (95%CI:0.17-0.84) for the CAEP study); however, it was not statistically significant in the CVC study sample.

## Discussion

This study presented the challenges associated with lacking a legal residency permit for Syrian refugees among two samples in Lebanon. Nearly all Syrian refugees reported not holding a legal residency permit in both samples. The lack of a legal residency permit was found to be associated with poor mental health status, lower uptake of COVID-19 vaccination, and lower access to healthcare services. While the literature has widely reported on the socio-economic and health challenges of Syrian refugees, including discrimination, restricted mobility, poverty, low education, and inadequate access to healthcare in general [2, 4–13], to our knowledge, limited literature has examined the interplay between legal residency and healthcare access and outcomes.

In both study populations, the proportion of Syrian refugees without a legal residency permit was comparably high, aligning with the 80% reported in recent literature and the latest reports in Lebanon [2, 4]. This highlights the host country policies that contribute to the legal vulnerability of refugees [4]. Consistent with previous studies, those with lower socio-economic status were less likely to have legal residency. This is likely due to the large financial burden and paperwork obligations for the annual residency permit renewal [4, 15, 16]. Furthermore, those without a legal residency permit were less likely to be employed. This finding is consistent with another analysis, which showed that Syrian refugees with legal residency had a higher likelihood of being be employed, potentially facilitated by their freedom of movement and access to work permits [19]. Conversely, the inability to obtain or renew legal residency may compel Syrians to engage in informal work, where they experience no job security and poor working conditions [6]. Gender disparities were also apparent, where a higher proportion of men possess legal residency documentation as compared to women [4].

The lack of a legal residency permit was associated with poorer mental health. This may be explained by direct and indirect pathways. For example, those without a permit may have an increased sense of insecurity and fear of arrest, detention, deportation, or protection risks including exploitation, all of which negatively impact mental health [4, 16]. Furthermore, an indirect association with mental health could be explained by being unable to gain employment, the consequent lack of means to provide for oneself, or having to seek employment within the informal sector due to lacking a legal residency permit.

In addition, legal residency status was associated with healthcare access, aligned with qualitative findings on Syrian refugees in Jordan [35]. This was observed in both of our studies, although statistical significance was not reached for the CVC study, likely due to study power. Lacking a legal residency permit may result in movement restrictions preventing refugees from travelling to healthcare centres [4], or alternatively they may not attend the centre itself due to fear of repercussions, as most healthcare facilities in Lebanon require a form of identification [24, 25]. There should, therefore, be a focus on improving healthcare access for refugees to be based on health need rather than residency status.

COVID-19 vaccine uptake was also lower among those lacking a legal residency permit, despite COVID-19 vaccines being available free of charge to all residents of Lebanon following registration through a government-generated platform. Fear of detention or deportation and mistrust of the Lebanese government are possible reasons why many Syrian refugees did not register [17]. Therefore, refugees must be targeted in vaccination campaigns, potentially through specific processes and accessible sites, and the needs of those without legal residency should be met in future emergency responses and pandemics.

This study is subject to some limitations. The study exposure was self-reported, which may be prone to recall bias; however, most refugees reported not having a legal residency permit. Furthermore, the CVC study sample is representative of older Syrian refugee beneficiaries from a single humanitarian organisation and not all Syrian refugees residing in Lebanon. It is likely comparable to older Syrian refugees receiving humanitarian assistance in Lebanon. The CAEP study includes all adult Syrian refugees in Sin El Fil who responded to the survey, which will be comparable to Syrian refugees residing in other suburban contexts in Lebanon.

## Conclusion

This study highlights the difficulties faced by Syrian refugees in Lebanon in securing legal residency permits and the resulting repercussions on health. It found that most Syrian refugees living in Lebanon reported not holding legal residency permits at the time of the data collection, and addresses an important knowledge gap in the association of legal residency with health outcomes. These findings highlight the need for actions to support the legal protection of refugees and enable equitable access to vaccination campaigns and health, and particularly mental health, services for this vulnerable population. More specifically, consideration for legal residency should be integrated into the recently launched National Mental Health Strategy in order to improve the mental health of Syrian refugees [36]. Moreover, access to healthcare services should be guaranteed regardless of legal residency status.

## Contributors

HG, SA and SJM conceptualized the parent studies; SJM, HG, SA, NS, BA and MER designed the surveys, contributed to study methodology and investigation, and oversaw the data collection. The formal analysis and literature search was conducted by MER. MER and SJM drafted the manuscript. SJM supervised MER, HS and NS throughout the project. MER, HG, BA, HS, NS, JD, SA and SJM contributed to the interpretation of the results, and have edited drafts and approved the final version of the article. MER, HS, HG and SJM had full access to and verified the raw data. SJM is the guarantor. All authors had access to the study data and had final responsibility for the decision to submit the manuscript for publication.

## Data sharing

The anonymized data can be obtained upon reasonable request from the Center for Research on Population and Health at the American University of Beirut.

## Acknowledgments

We would like to thank the “Community Action for Equity in Pandemic preparedness and control” CAEP Study Group for supporting the CAEP study work (Nada M. Melhem, Aline Germani, Fadi El-Jardali). We acknowledge BOT (Bridge. Outsource. Transform) for their assistance in collecting the data required for the success of this study and the study participants for their participation.

