## Supplementary material for "Legal residency status and its relationship with health indicators among Syrian refugees in Lebanon: a nested cross-sectional study"

**Supplementary Materials**

Supplementary Figure 1. Flow diagram of the study population of older Syrian refugees from all areas of Lebanon who completed the survey at wave 1 for Study A.

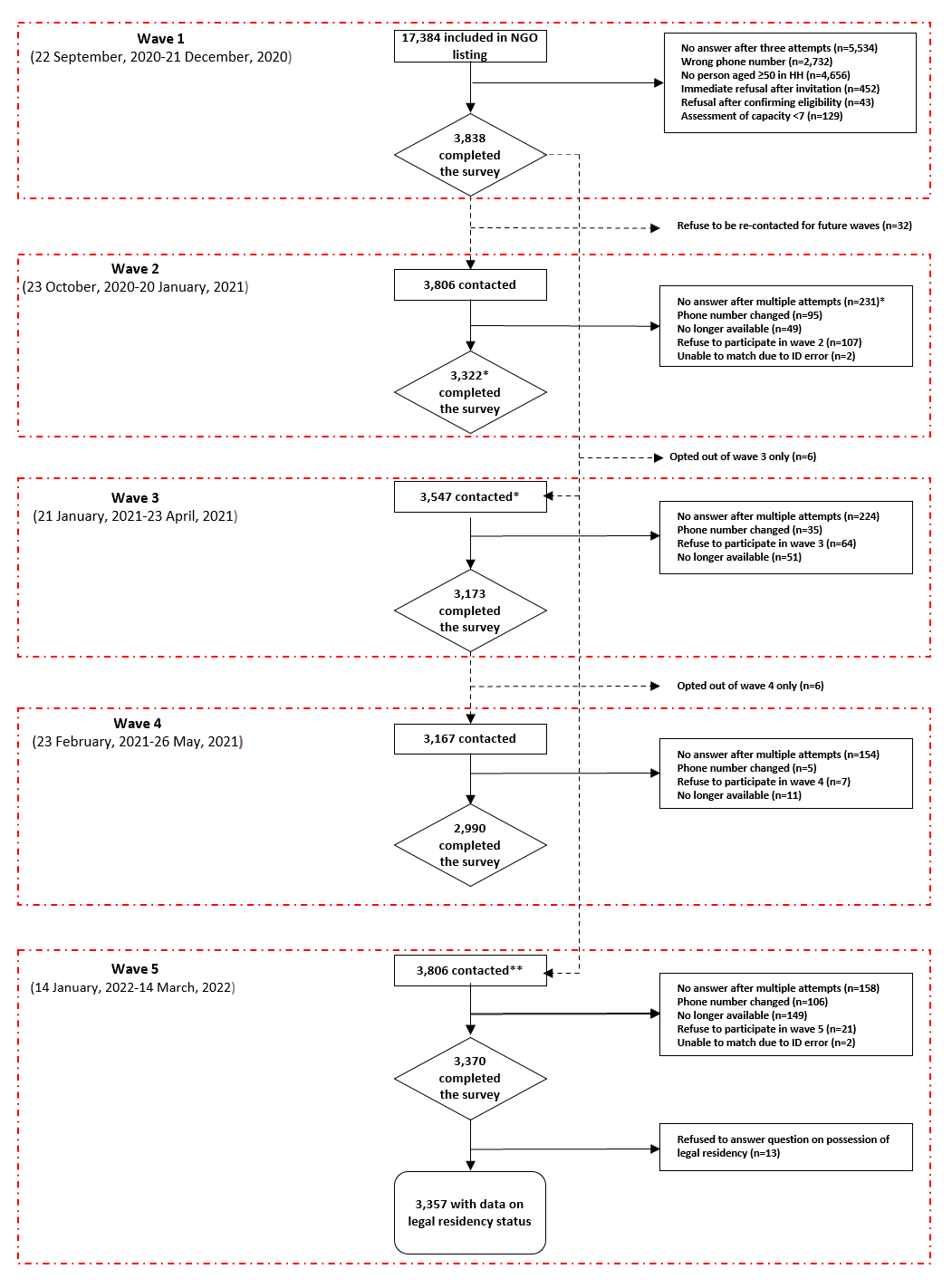

**Wave 3 included all participants who participated in wave two, and those who didn’t answer after multiple attempts at wave 2. **All participants who provided consent in wave one were re-contacted at wave five.*

Supplementary Figure 2. Flow diagram of the study population of adult Syrian refugees residing in a district of Beirut, Lebanon who completed the survey at wave 1 for Study B.
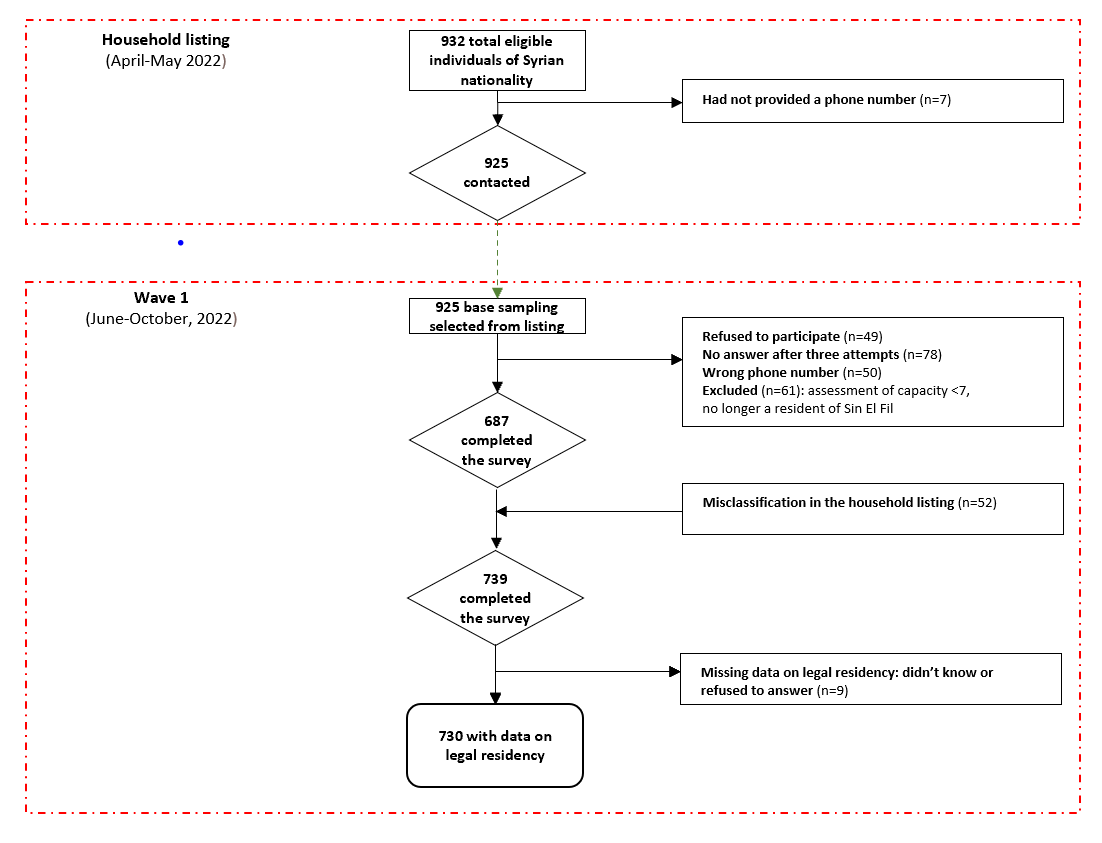

Supplementary Figure 3. Directed Acyclic Graph for the association between legal residency status and each health outcome: mental health, COVID-19 vaccination, primary healthcare access and disease management/medication access.

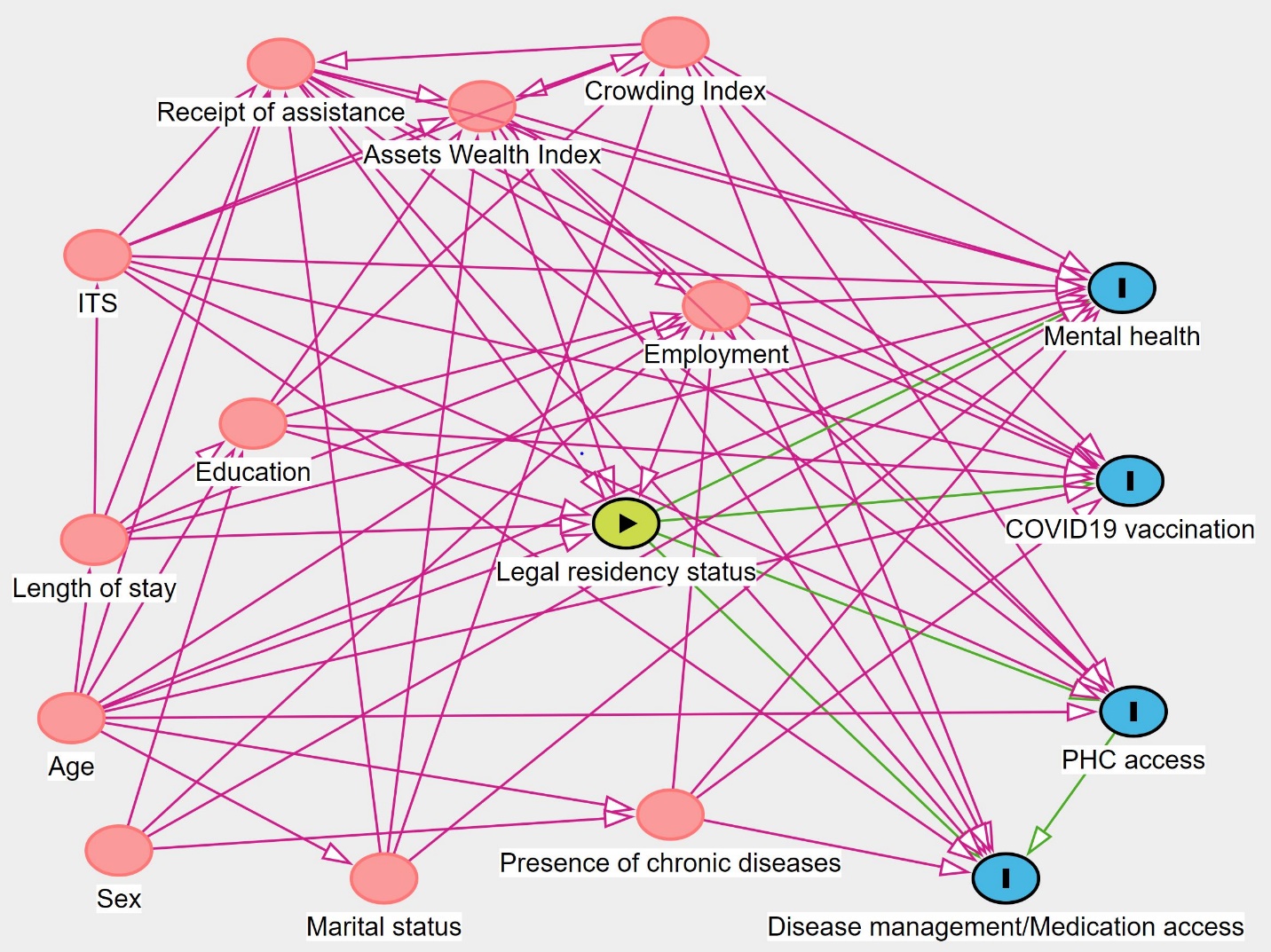

*Minimal sufficient adjustment set for each health outcome included age, length of stay in Lebanon, education, employment, wealth index, and receipt of assistance.*

*PHC = primary healthcare*
